# Experimental Methods of Investigating Airborne Indoor Virus-Transmissions Adapted to Several Ventilation Measures

**DOI:** 10.1101/2022.01.10.22268982

**Authors:** Lukas Siebler, Maurizio Calandri, Torben Rathje, Konstantinos Stergiaropoulos

**Author notes:** Contributing authors.

## Abstract

This study introduces a principle, which unifies two experimental methods for airborne indoor virus-transmissions adapted to several ventilation measures. A first-time comparison of mechanical/natural ventilation and air purifiers with regard to infection risks is achieved. Effortful computational fluid dynamics demand detailed boundary conditions for accurate calculations of indoor airflows, which are often unknown. Hence a suitable, simple and generalized experimental set up for identifying the spatial and temporal infection risk for different ventilation measures is required. A trace gas method is suitable for mechanical and natural ventilation with outdoor air exchange. For an accurate assessment of air purifiers based on filtration a surrogate particle method is appropriate. The release of a controlled rate of either trace gas or particles simulates an infectious person releasing virus material. Surrounding substance concentration measurements identify the neighborhood exposure. One key aspect of the study is to prove that the requirement of concordant results of both methods is fulfilled. This is the only way to ensure that the comparison of different ventilation measures described above is reliable. Two examples (a two person office, several classrooms) show how practical both methods are and how the principle is applicable for different types and sizes of rooms.

---

**Table 1.**
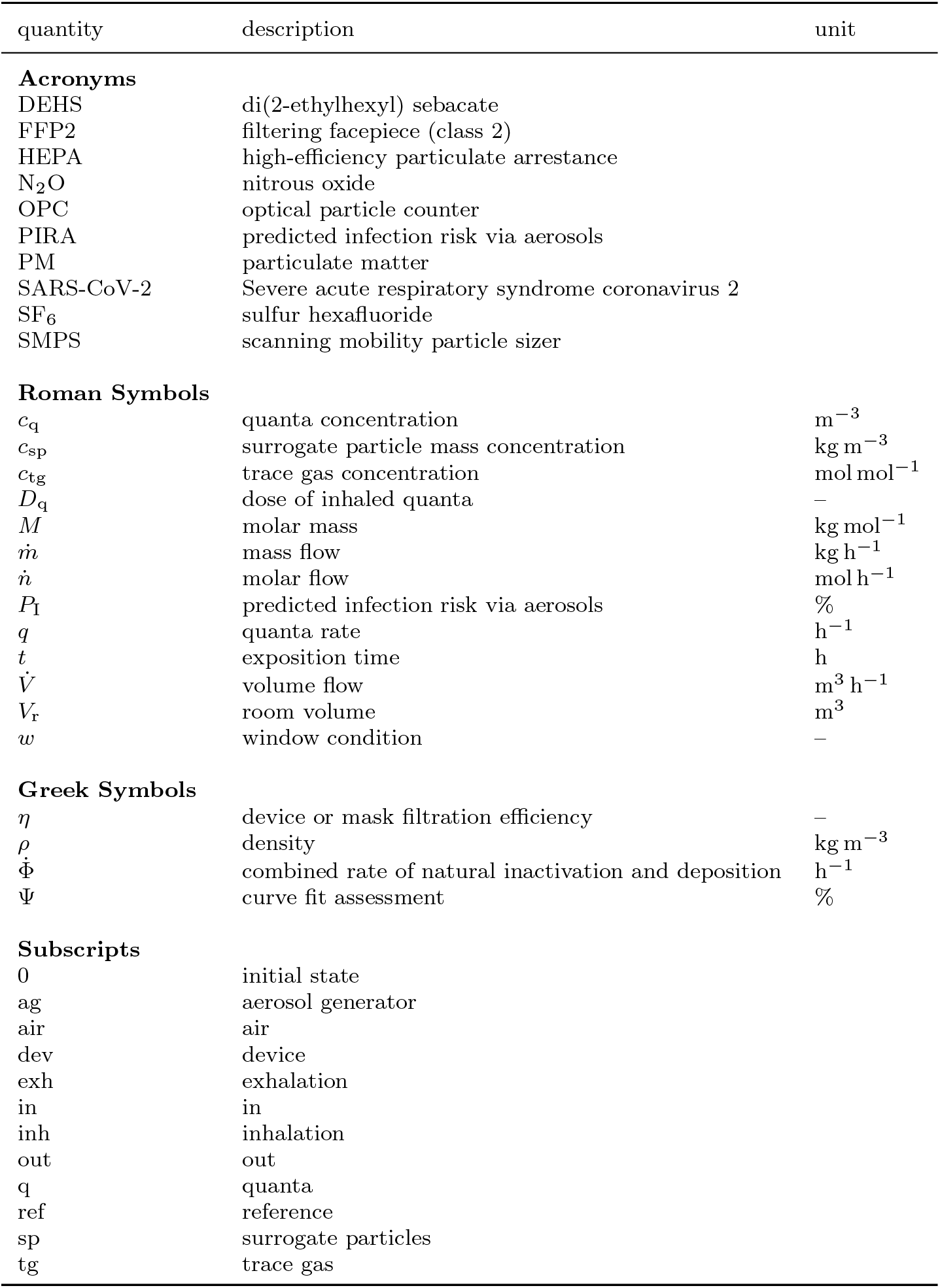
nomenclature

## 1 Introduction

The ongoing pandemic teaches us that the implementation of appropriate ventilation measures can significantly reduce indoor infection risks.

For fast and simple assessments many calculation models regarding the infection risk of SARS-CoV-2 in indoor environments exist [1–6]. Using idealised assumptions (e.g. ideal mixed ventilation) the accuracy decreases with the size of the room. Even for small to medium scales, these may not resolve aerosol dispersion appropriately. Displacement ventilation concepts, often applied in large halls, impede estimations of virus transmission to neighbors. In ideal theory there would occur only unobjectionable vertical buoyancy flows. In reality, however, these are superimposed by horizontal flows due to disturbance effects (e.g. cold walls and leaky doors), which are critical regarding neighbor infection risks. If the relevant boundary conditions for these disturbing influences are unknown, the estimation is regarded even more critical.

For individual considerations and higher accuracies, experimental studies examine the effects of ventilation measures on airborne infections. Some of them deal with a real virus load of the indoor air with actually present infectious persons, without controlled boundary conditions [7]. In [8], trace gas measurements and flow simulations are used to simulate an actual infection scenario in order to reproduce transmission paths in detail. Transient and absolute considerations of virus loads are omitted in this context. Other research studies examine the effects of ventilation measures, but do not take into account the transfer of substitutes to viruses and thus the actual infection risk [9]. As a result, only relative statements on the risk reduction are possible.

In general, ventilation measures can be divided into filtering recirculation methods (e.g. air cleaning) and ventilation principles with outside air exchange (e.g. natural/mechanical ventilation). However, there are no published studies on the comparative assessment of both measures with regard to the infection risk.

Previous experimental research is not yet suitable for a holistic individual assessment of the infection risk. Therefore, a comprehensive and standardized measurement procedure for any airborne transmitted disease is required. In this paper, two consistent measurement methods are presented, which allow to determine spatially and temporally resolved infection risks in rooms accurately. For the first time, the two consistent approaches can be used to evaluate both filtering recirculation devices and ventilation principles with outdoor air exchange.

## 2 Theory of airborne virus infections

In 1955, the first approach of assessing general infection risks was developed, which was improved and specified in 1978 on the disease measles [10, 11]. The authors introduced the dose of inhaled quanta *D*_q_ as indicator whether an infection occurs. Taking into account the efficiency of masks it can be calculated as:

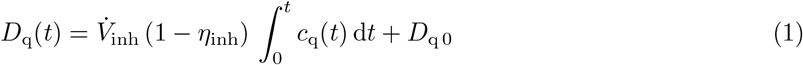

with 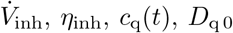 as inhalation volume flow, mask filtration efficiency for inhaling, quanta concentration at time *t* and the initial value of *D*_q_ respectively.

The approach of *Wells et al*. and *Riley et al*. allows the computation of the predicted infection risk via aerosols (PIRA), labeled as *P*_I_:

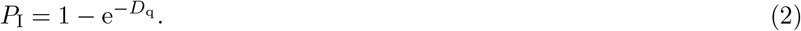

### Continous ventilation

Rooms in non-residential buildings are commonly equipped with continuously operating ventilation systems. The following figure shows the transient course of quanta concentration and the resulting PIRA over time (fig. 1) [12].

**Fig. 1.**
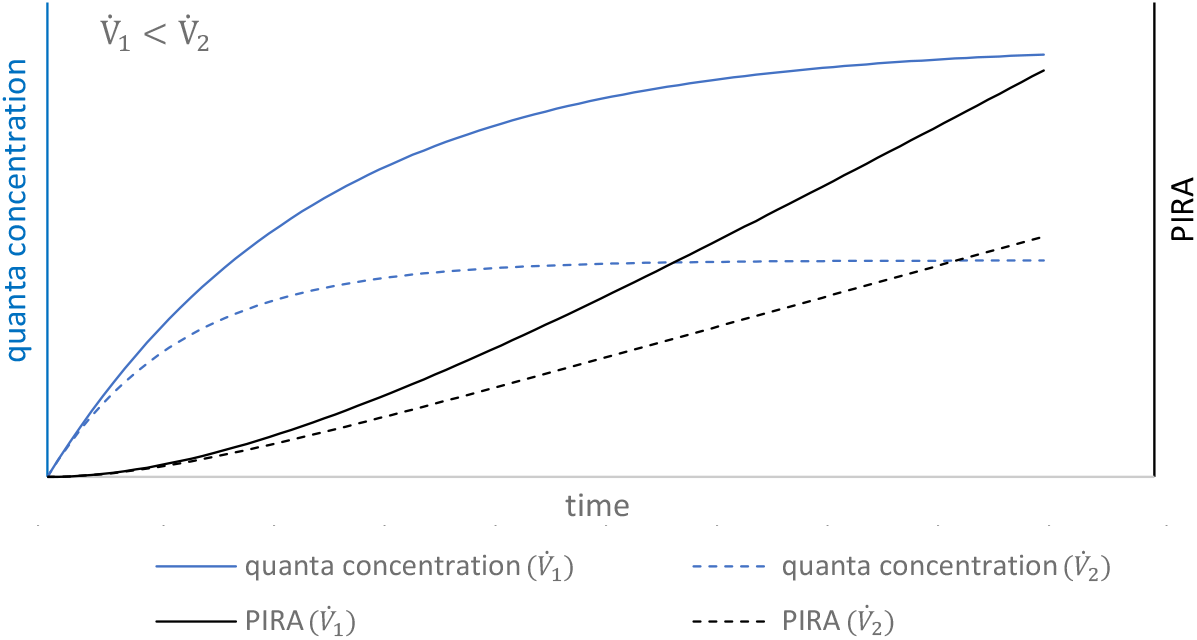
quanta concentration and PIRA course for different device volume flows

For this continuously ventilation the blue curves describe the solution of an inhomogeneous differential equation of first order and the black curves the resulting PIRA for two different volume flows respectively.

The shape of these theoretical curves is similar, when replacing a ventilation device by an air purifier. Only the absolute values can change because of the degree of particle separation of the device. Furthermore, for an air purifier only one undesired short circuit effect exists (supply to extract air) instead of two (supply to extract air and exhaust to outdoor air) for a ventilation device.

Equation (3) shows the differential equation for the quanta concentration for a continuous ventilation:

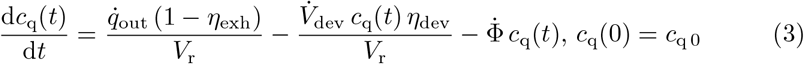

with 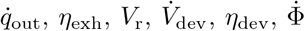 as quanta rate (output), mask filtration efficiency for exhaling, room volume, volume flow of device and device efficiency (filtration ratio for air purifiers, exhaust air to outdoor air exchange for ventilation systems) and combined rate of natural inactivation and deposition of existing viruses respectively.

The differential equation (3) consists of three different terms. The first one is based on the continuous release of quanta of an infectious person, the second is the quanta removal of the ventilation according to the current quanta concentration and the third is a combined rate of natural inactivation and deposition of existing viruses [13–16]. The last term can be described and modelled well in theory, but in practice it is challenging to include it in experimental settings. In case of zero initial concentration this equation can be solved analytically resulting in:

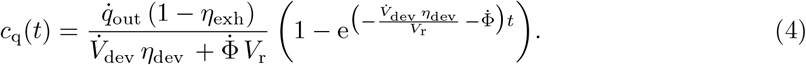

Besides the exposition time the volume flow has the most relevant impact on the infection risk. The higher it is adjusted the lower the equilibrium level of the concentration and therefore the infection risk (fig. 1).

### Periodic ventilation

For a periodic natural ventilation, the course of quanta concentrations show different sections depending on the window opening strategy. There is a switch between a differential equation for closed and for opened windows. This can either be solved with an analytical [17] or a numerical approach (less effort). The following plot (fig. 2) shows the quanta concentration over time for a periodic strategy of natural ventilation depending on different cross-section areas of the windows and resulting volume flows [18]. Without the considered deposition and natural inactivation the curve *no window* and the sections of closed windows would become straight lines. Besides a homogeneous substance distribution (at any position at any time) it is assumed that the wind velocity and the outside temperature are constant and that the room temperature is hold at a constant value by an ideal controller of the heating system.

**Fig. 2.**
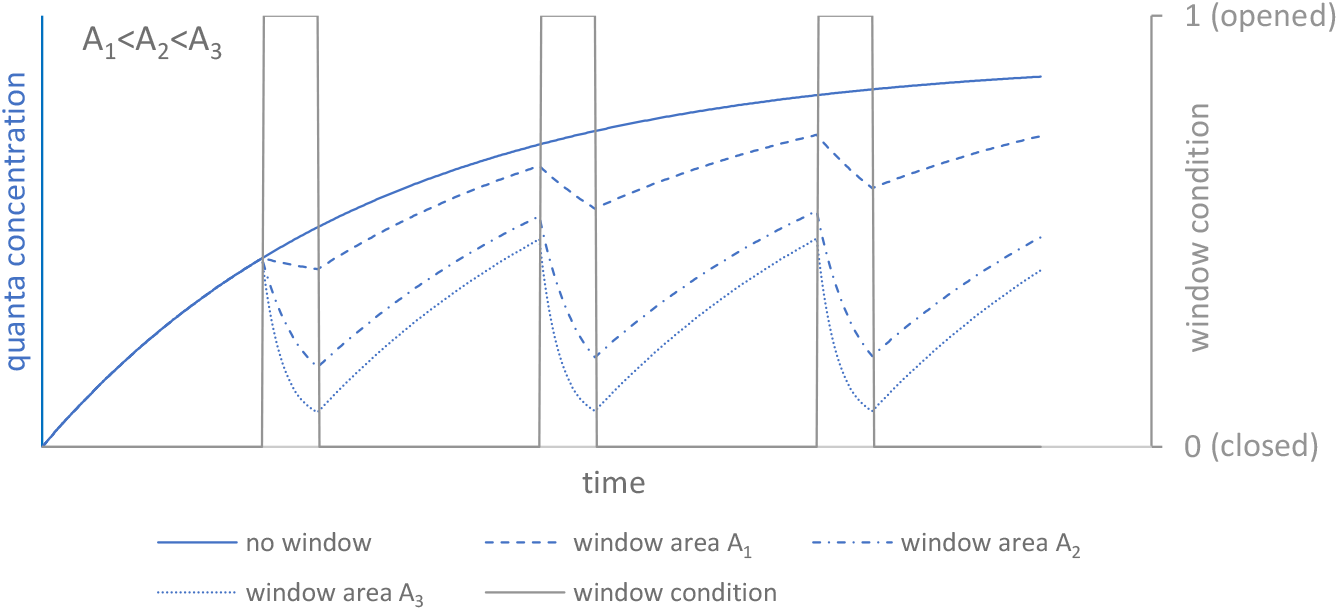
quanta concentration course for periodic ventilation depending on different opened window areas

The step-wise differential equation for the transient concentration and the resulting *D*_q_ is shown in (5) and (6):

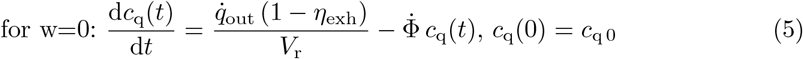

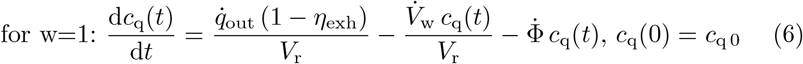

with 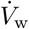 as window volume flow. Using (1) and (2) the dose of inhaled quanta and finally PIRA can be calculated. The higher the window area and therefore the resulting periodic airflow, the lower the dose of inhaled quanta and thereby the infection risk.

With these assumptions it is possible to estimate infection risks using ventilation devices, air purifiers and natural ventilation under mixed ventilation conditions. Whereas experimental methods of substance dispersion concerning the airborne transmission of virus infections enable accurate spatially and temporally resolved results for any ventilation principle even for unknown boundary conditions.

## 3 Experimental Methods

Airborne virus transmission is mostly dealing with particles below 5 µm of size [19, 20]. These particles are meant to have negligible sink velocities and follow airflows exactly [21]. A common method investigating airflows quantitatively is releasing substances (which also follow the airflow) and measuring their concentrations. In order to evaluate the totality of the ventilation measures suitable substances are trace gas and surrogate particles. For scenarios with outside air exchange only trace gas is appropriate because of entering particles from the outside, which would falsify particle measurements. For recirculation devices based on filtration, however, trace gas is inapplicable since it is not filtered and therefore no device effect can be measured. For this reason, two different methods are essential.

### 3.1 Trace gas method

The focus of previous indoor air investigations using trace gas, also conducted at the University of Stuttgart, has mostly been on evaluating the ventilation effectiveness rather than infection risks [22]. The method is based on the emission of trace gas, which is not present in the natural surrounding and whose concentration can be measured by infrared spectrometers (e.g. N_2_O or SF_6_). First the room needs to be freed from trace gas from eventual previous measurements. By using a mass flow controller with gas specific adapted properties a constant and continuous mass flow of the trace gas is possible.

Along with the measured trace gas concentration, the quanta rate (output) and the respiratory rate the following theoretical post processing leads to a calculation of the infection risk of a certain disease.

For trace gas, which is transported in a similar way to airborne particles and viruses (all below 5 µm) [23] the following approach without mask filtration efficiency is assumed:

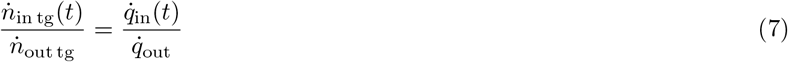

with 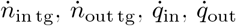 as (fictitious) molar flow for trace gas (input), molar flow for trace gas (output), quanta rate (input) and quanta rate (output) respectively.

From the (time dependent) quanta rate (input) 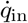 the dose of inhaled quanta *D*_q_ is determined by integration over time (assumption: *D*_q 0_ = 0, *c*_q 0_ = 0):

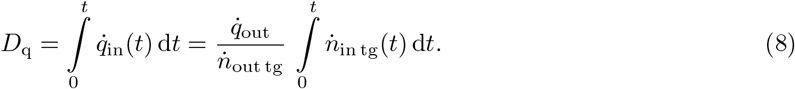

The molar flow for (fictitious) trace gas (input) is calculated by using 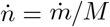 and the assumption *c*_tg_(*t*) ≪ 1:

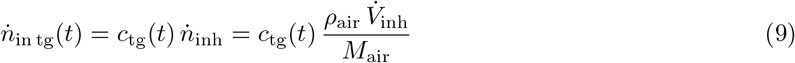

with 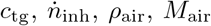 as measured trace gas concentration, inhalation molar flow, density and molar mass of air respectively (note: conversion from ppm in mol mol^*-*1^ might be necessary).

The resulting equation for the dose of inhaled quanta *D*_q_, considering mask filtration efficiency (see [24, 25]), is:

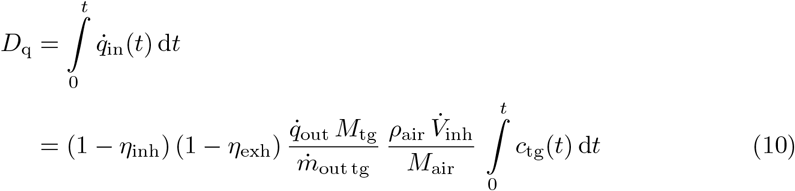

with 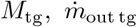 as molar mass and mass flow of trace gas (output) respectively.

A numerical integration of the concentration of trace gas over time *c*_tg_(*t*) (e.g. via trapezoidal rule) allows the transfer into measurement data via equation (2) into an infection risk for scenarios with outdoor air exchange.

### 3.2 Surrogate particles method

To evaluate the function of an air purifier based on filtration an alternative method is required, because trace gas can not be filtered. For this approach the room needs to be freed from particles by the air purifier first. From then on only particles that are actually released by particle generators are measured. A suitable substance for particles is Di-Ethylhexyl-Sebacat (DEHS). Besides the advantage that its particles keep the same size over time because of very low evaporation rates, the measured size distribution is similar to the exhaled particles of humans [20, 26].

Any other possible particle sources (even persons) should be removed from the room of investigation or kept emitting as low as possible. Using several optical particle counter (OPC) or Scanning mobility particle Sizer (SMPS) on different positions allow a spatial and temporal measurement of the number concentration and size distribution of particles, which are visible for these instruments.

The measured surrogate particle concentration, the quanta rate (input) and the respiratory rate lead to the calculation of the infection risk for a room equipped with an air purifier.

Analogue to the trace gas method for surrogate particles the following approach without mask filtration efficiency is assumed:

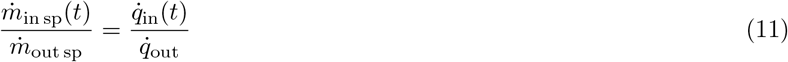

with 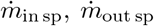 as mass flow for surrogate particles input (fictitious) and output respectively.

Similar to the trace gas method the dose of inhaled quanta *D*_q_ is (assumption: *D*_q 0_ = 0, *c*_q 0_ = 0):

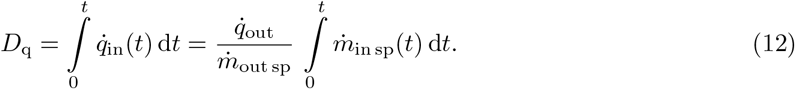

Substituting the assumed constant mass flow (output) of surrogate particles 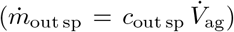 and a fictitious mass inhalation rate of surrogate particles 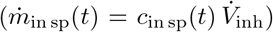 in (12) (considering mask filtration efficiency see [24, 25]) results in:

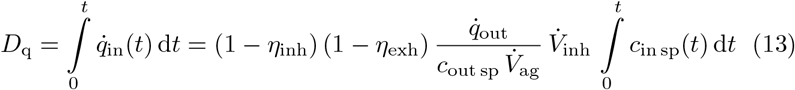

with 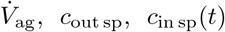 as volume flow of aerosol generator, mass concentration of surrogate particles (output and input) respectively.

With regard to suspected agglomeration effects a mass range up to 10 µm is suggested for higher levels of accuracy (see also section 6). This can be determined by cumulating mass concentrations of each size fraction by calculating their volumes and their weight with the density of the specific particle substance.

## 4 Comparison between both methods

One key aspect of the study is to prove that both methods are concordant. Hence, a comparison between both measuring methods for substance dispersion is essential. *Bivolarova et al*. compare trace gases with different particle sizes on the basis of steady-state observations and decay curves of substance concentrations. A temporal resolution of these concentrations from the initial to the steady state - i.e. the dispersion of the substances - as well as a transfer to an infectious event is not focused here. [27]

However, depending on the infectiousness of a disease and the exposure time, the transient dispersion is relevant to an infection process. In addition, the relation of substance release rates (emission) to emerging concentration profiles (immission) is not yet examined and thus a transfer to a fictitious unit such as quanta is not possible. For these reasons, a comparison is made with respect to transient quanta concentration courses in an airflow visualization laboratory with controlled boundary conditions. In order to ensure that almost no outdoor particles enter the room a HEPA 14 filter was integrated into the duct of supply air. Otherwise, in addition to the released particles, those from outside would also be measured and therefore corrupts the experimental data. A simultaneous release and measuring of particles and trace gas at the same position is expected to result in the same dispersion of both substances. In this case the calculated course of the curves of quanta concentrations for both substances is supposed to be similar in all measuring positions. This experimental set up is shown in figure 3.

**Fig. 3.**
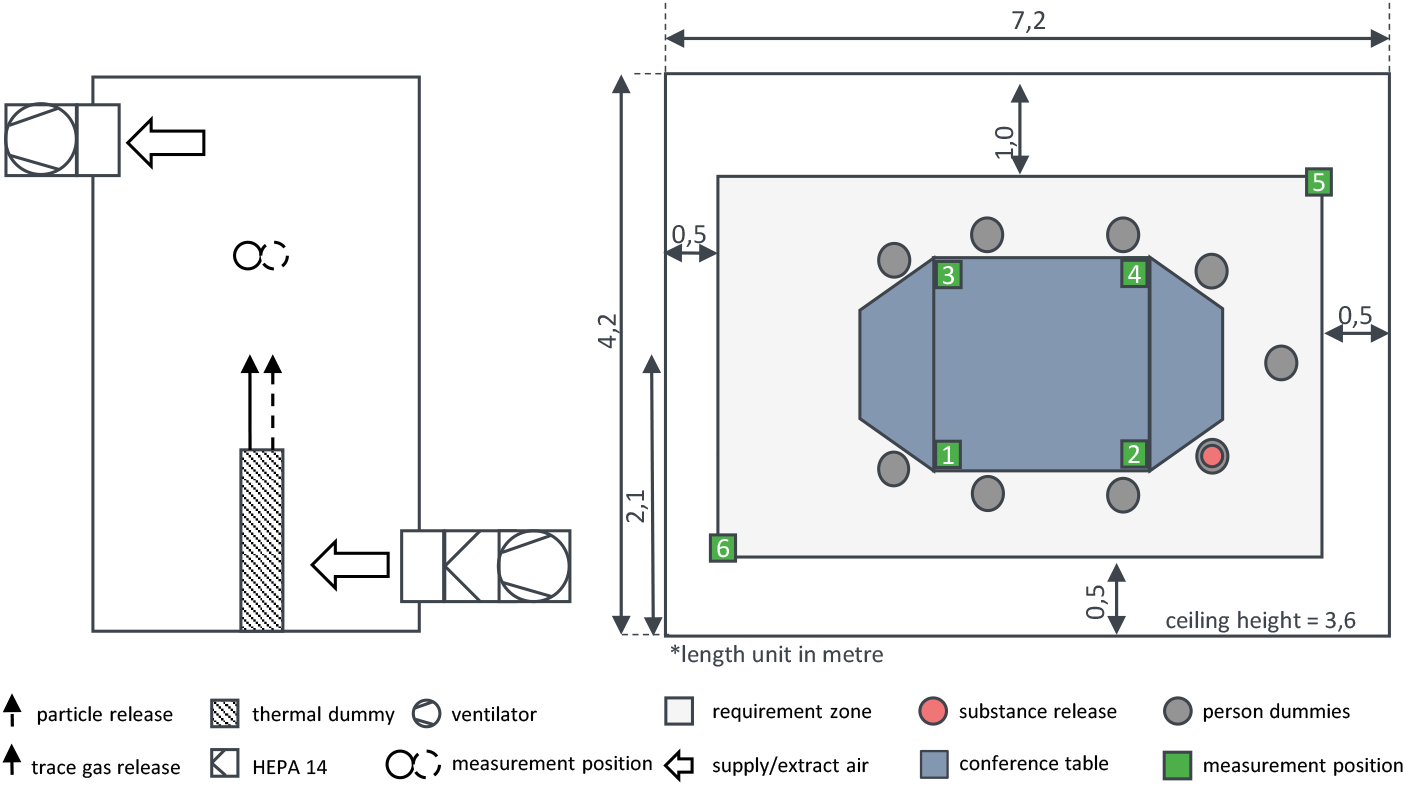
set up to compare both methods (left) and top view of the conference room (right)

**Table 2.** relevant parameters of the comparative measurement

| quantity | value | quantity | value |
| --- | --- | --- | --- |
| room volume | 109 m<sup>3</sup> | volume flow | 1250 m<sup>3</sup> h<sup>-1</sup> |
| quanta rate (input) | 46.5 h<sup>-1</sup> | exposition time | 1 h |
| medium | SF<sub>6</sub>/DEHS | number of persons | 9 |

In figure 4 the result at a single measuring position (best fit) in the conference room is shown. Over the entire measurement period both quanta concentration courses are similar. Therefore it is expected that both methods are suitable and can be applied for different ventilation cases. Also worth emphasizing is the small discrepancy between both experimental curves compared to the theoretical one under steady state conditions. This is due to the possibility of accurate volume flow measurements (correlation to steady state concentration) in the laboratory. However, during the transient course, a discrepancy is generally observed, since substances in reality need time to disperse to a certain position.

**Fig. 4.**
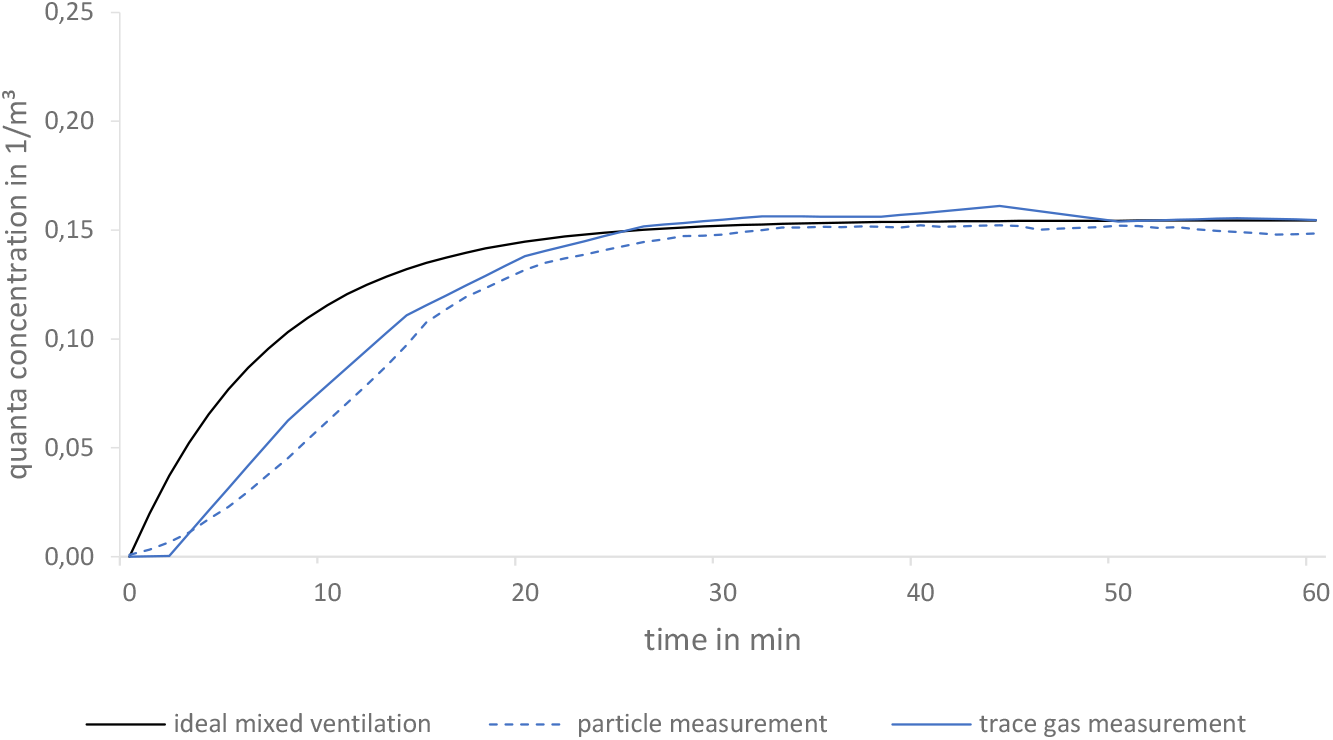
comparison between particle and trace gas measurement as a function of quanta concentration over time exemplified in a conference room (position 3)

## 5 Experimental studies

In the following section it is shown that both methods are practical. The measured scenarios go from small to large scale and contain the three main ventilation types: a two person office (air purifier), classroom (natural and mechanical ventilation, air purifier) and the Opera Stuttgart in Germany (mechanical ventilation). Except of the opera the investigated example scenarios are shown in detail.

Besides supply and extract air effects indoor airflows are influenced by sub- and over-tempered surfaces (e.g. thermal sources). For an appropriate simulation of human heat outputs for a certain activity rate (low activity: 75 W) [28] thermal dummies with a surface area of an average human (1.8 m^2^) [29] are recommended. Figure 5 shows a professional thermal person dummy and also a low cost method for substituting many persons in large rooms (e.g. Stuttgart Drama Theatre).

**Fig. 5.**
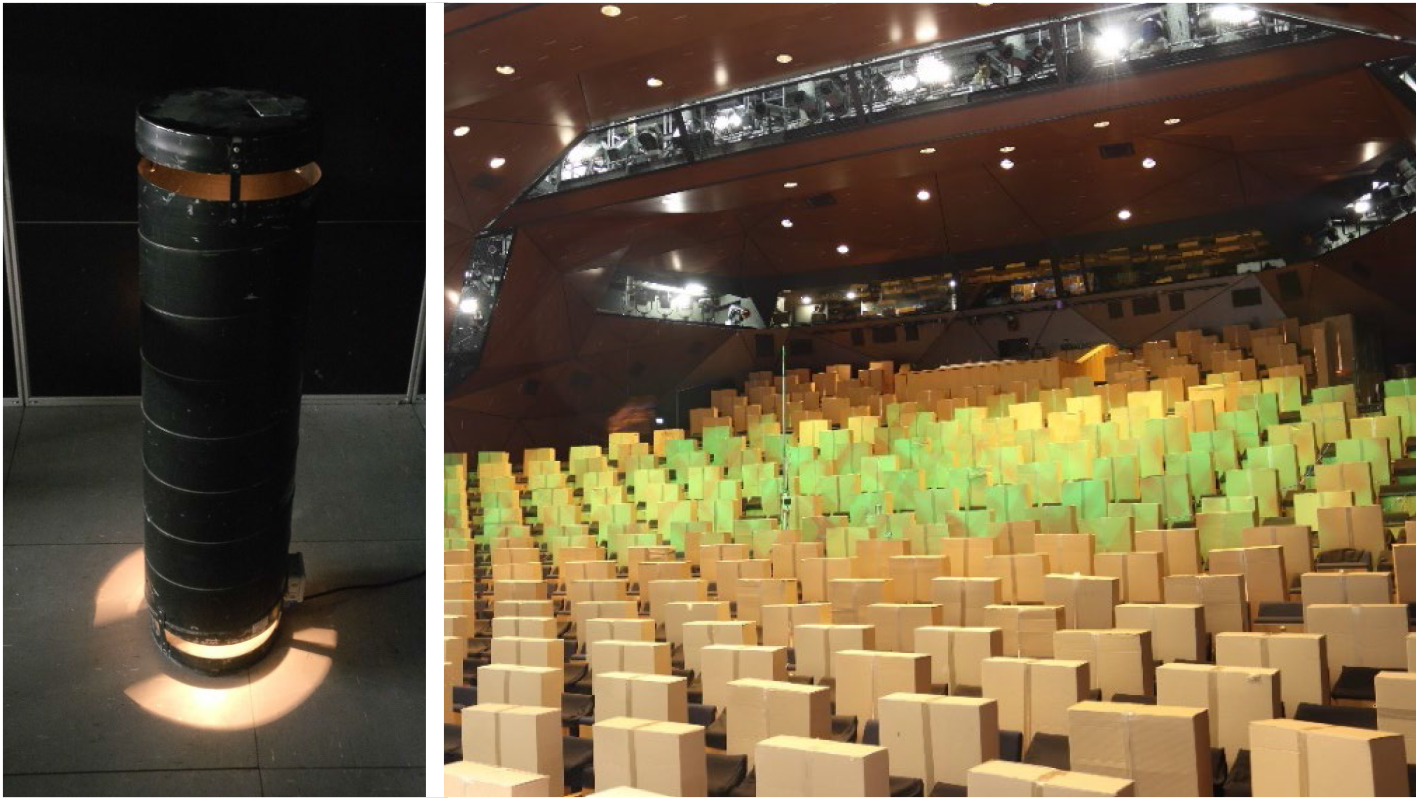
professional thermal person dummy (left), low cost variants (cartons with lightning bulbs inside) in Stuttgart Drama Theatre (right)

### 5.1 Spatially resolved measurements in a laboratory

Under reproducible boundary conditions in an air visualisation laboratory, the different air purifiers have been investigated and assessed. The focus lies on the particle removal effectiveness of the device in a two person office. Thereby one of these persons is assumed to be infectious. Figure 6 shows a photo and the top view of the experimental set up.

**Fig. 6.**
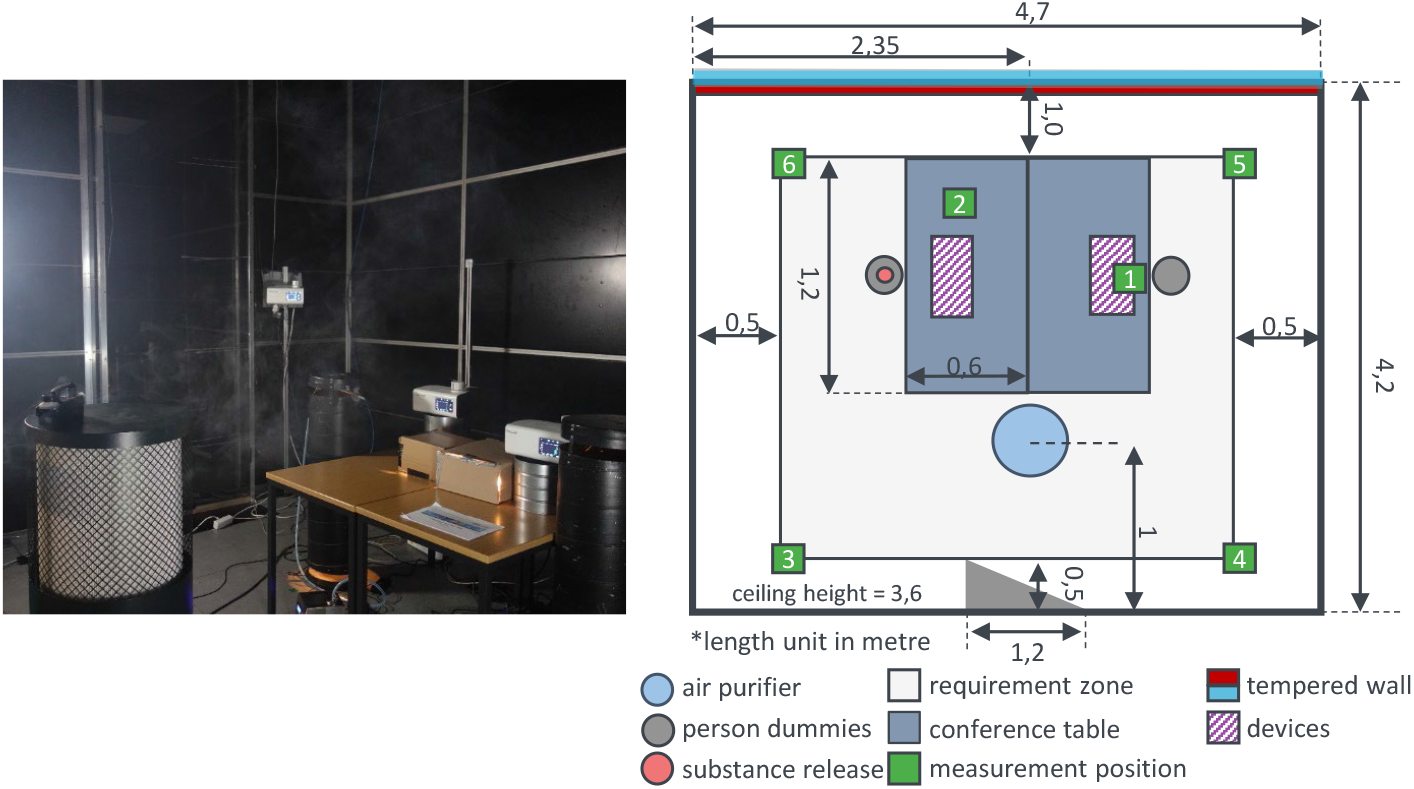
photo (left) and the top view (right) of the experimental set up of a two person office with an air purifiers in an air visualisation laboratory

**Table 3.** relevant parameters of the experiments concerning the effectiveness of air purifiers.

| quantity | value | quantity | value |
| --- | --- | --- | --- |
| room volume | 71 m<sup>3</sup> | volume flow | 900 m<sup>3</sup> h<sup>-1</sup> |
| quanta rate (output) | 46.5 h<sup>-1</sup> | exposition time | 1 h |
| medium | DEHS | filtration class | HEPA 14 |
| number of persons | 2 |  |  |

The temporally-resolved particle concentrations by the OPCs allow a computation of the infection risk (PIRA) at certain positions in a room using equation (13). An exemplary measurement result is shown in figure 7.

**Fig. 7.**
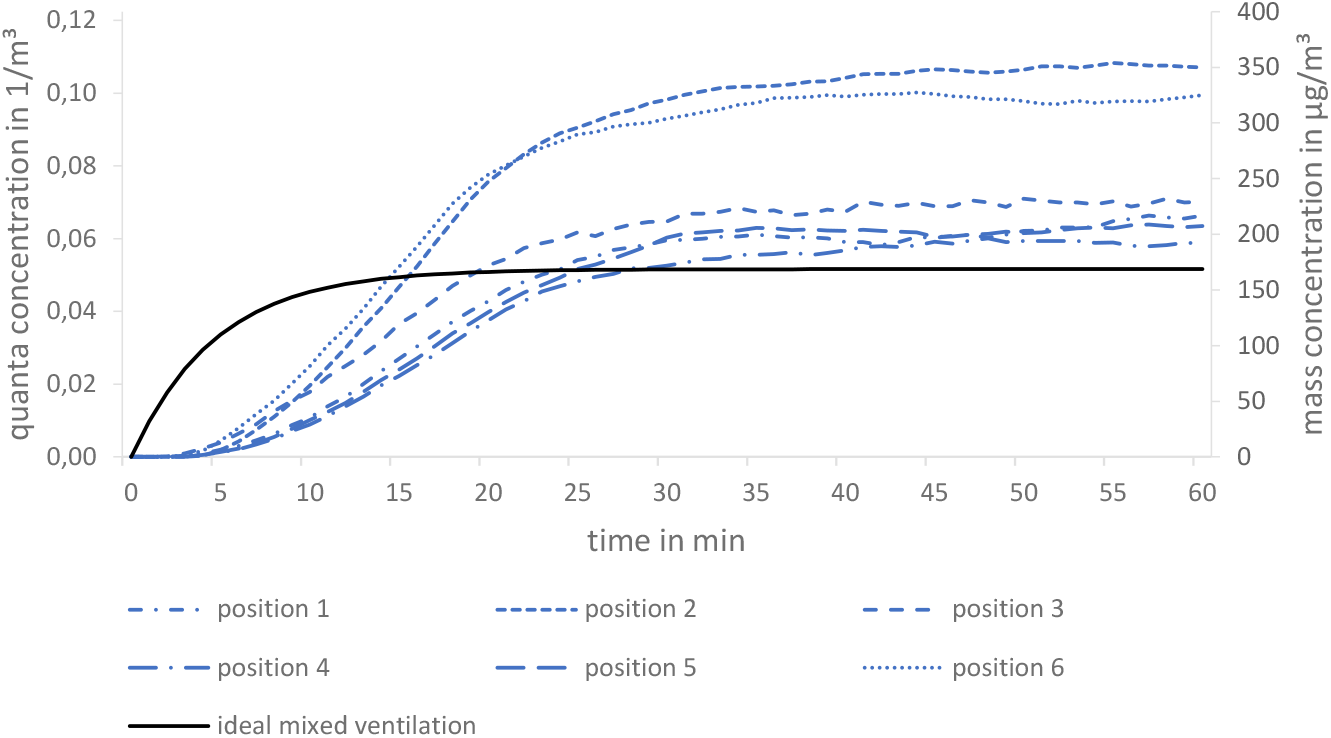
theoretical (ideal mixed air) and measured quanta concentration (left axis) and their related mass concentration (right axis) of six positions in a two-person office

The deviations of the individual positions among each other show a significant benefit of a spatial view. Moreover, quanta concentrations in figure 7 do not only mismatch the ideal mixed ventilation in the transient range but also in the steady state. The volume flow rates stated by the manufacturers of air purifiers (used for this calculation) seem to be less accurate than own measurements of a mechanical ventilation in an air duct (see fig. 4). In this case for a duration of 1 hour the overall infection risk for a medium spreader by Peng et al. [30] using equation (2) results in a range from 2.2 % (position 4) and 4.0 % (position 2).

### 5.2 Comparison of ventilation measures in classrooms

Further investigations of ventilation measures in classrooms were conducted, to assess the effectiveness of periodic window ventilation, decentralised ventilation systems and air purifiers regarding infection risk and thermal comfort. For this study, it is required that both methods (trace gas and surrogate particles) perform similarly. Figure 8 shows the set up and the three ventilation measures exemplarily.

**Fig. 8.**
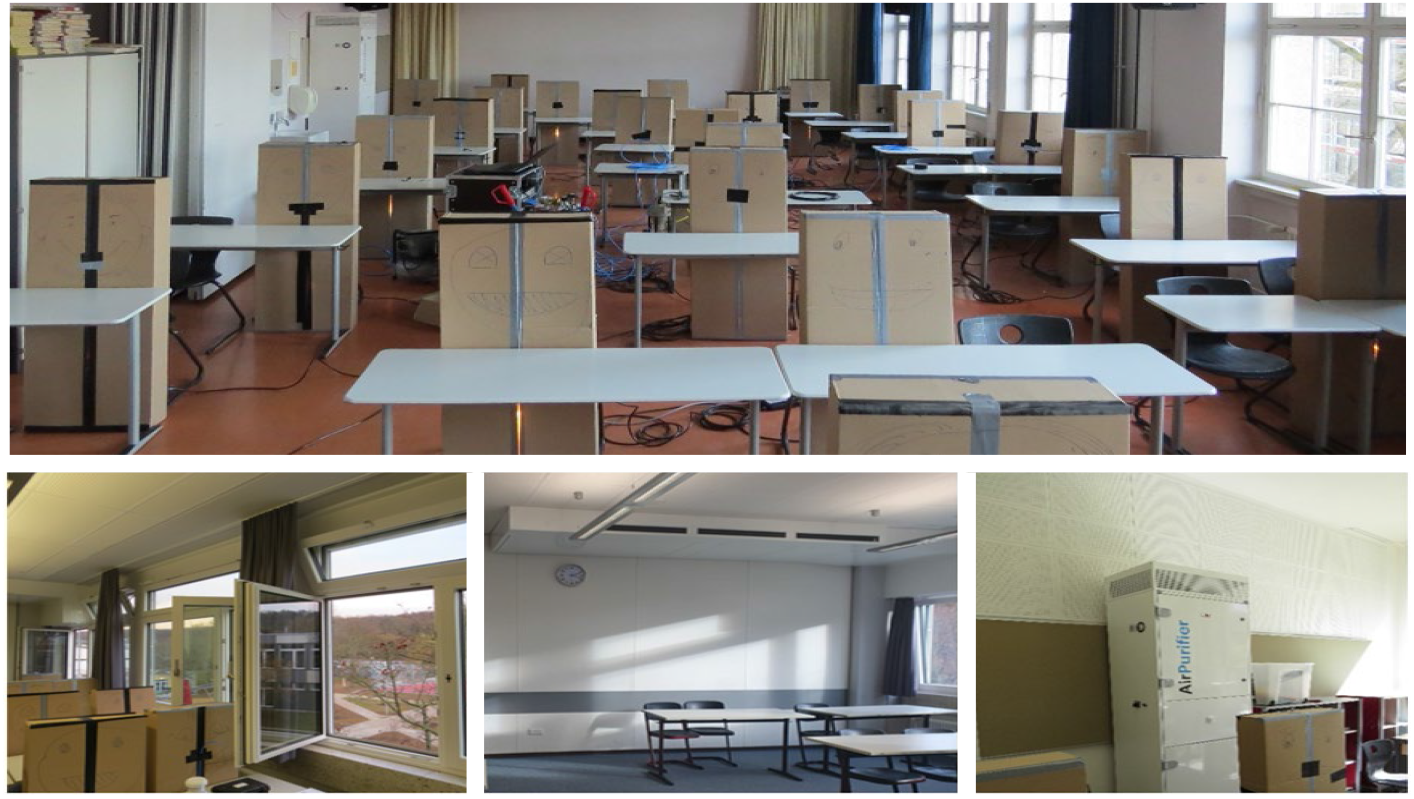
experimental set up (top), exemplary window ventilation (bottom left), exemplary decentralised ventilation system (bottom middle), exemplary air purifier (bottom right)

**Table 4.** relevant parameters of the experiments in classrooms

| quantity | value | quantity | value |
| --- | --- | --- | --- |
| room volume | 180...400 m<sup>3</sup> | volume flow | 500...1250 m<sup>3</sup> h<sup>-1</sup> |
| quanta rate (output) | 46.5 h<sup>-1</sup> | exposition time | 1.5 h |
| medium | SF<sub>6</sub>/DEHS | number of persons | 17...30 |

Due to changing boundary conditions during this study (compared to reproducible conditions in the laboratory), measurements were taken in many different rooms over several days. The results of the three investigated measures are illustrated in two plots in figure 9 and 10. They show the mean infection risk depending on the occurring volume flow for an assumed medium-spreader [30] and an exposition time of 1.5 h. For the periodic window ventilation (in this case 20/5/20: 20 min opened, 5 min closed) the current airflows are calculated depending on weather and window data [18]. The plots contain also the results for ideal mixed ventilations, which agree well with the measured data. Moreover, it should be emphasized that the comparison of air purifiers based on filtration and ventilation systems with air exchange (fig. 10) is only reliable because both measurement methods of the underlying principle match well (section 4).

**Fig. 9.**
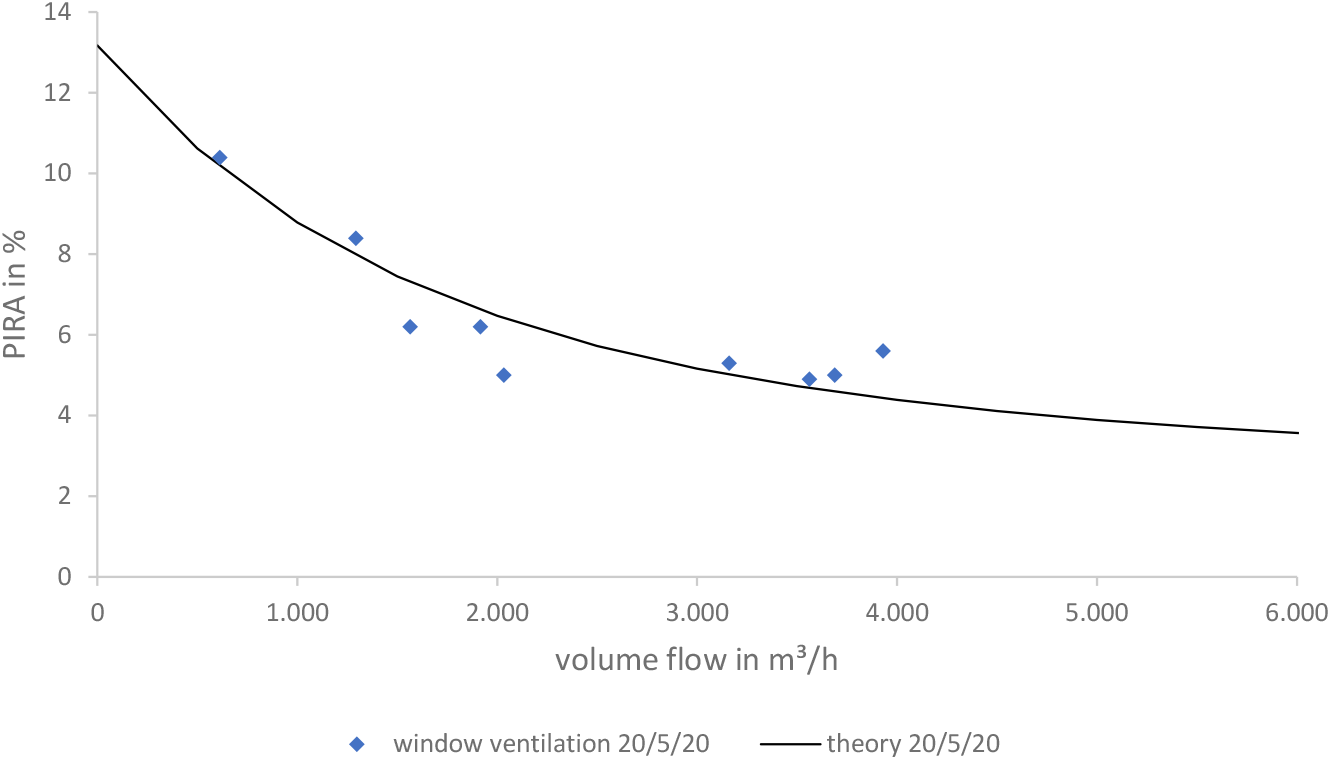
infection risk over occurring volume flow for periodic window ventilation

**Fig. 10.**
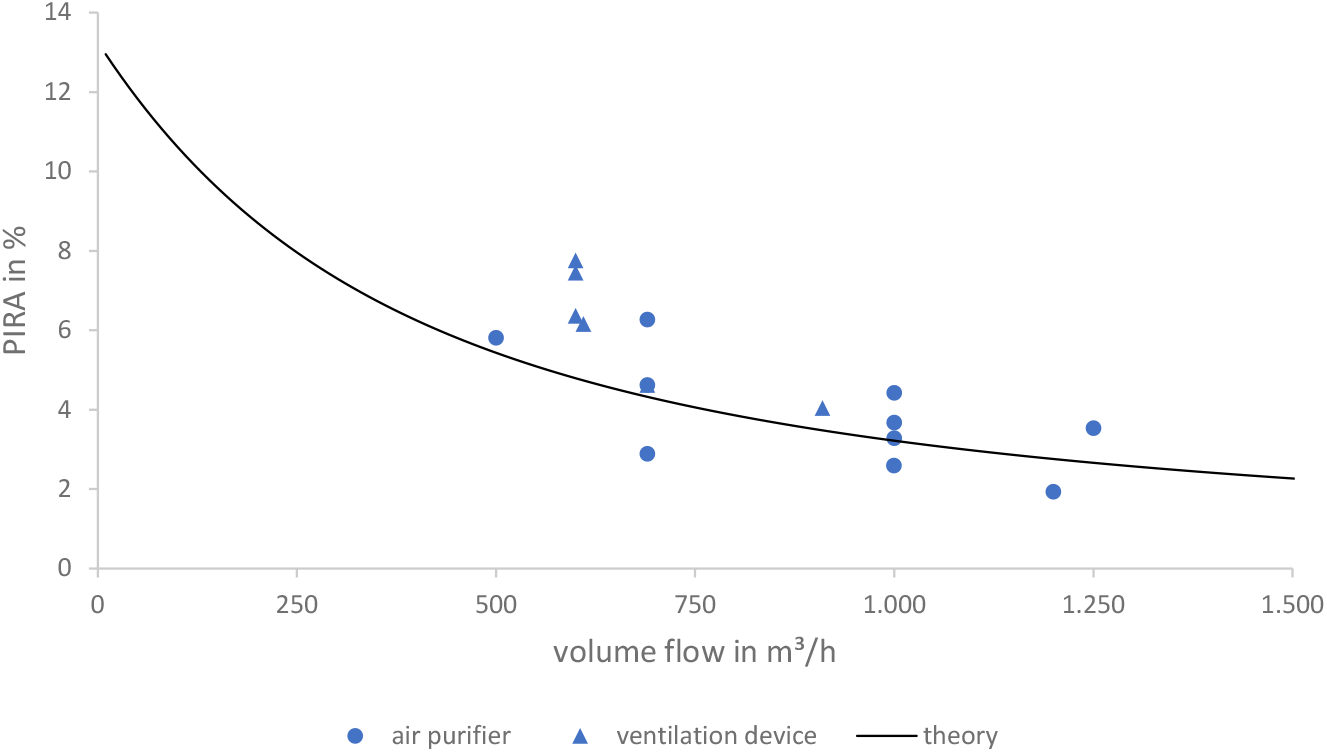
infection risk over continuously operating ventilation systems or air purifiers

## 6 Discussion

The behaviour of particles in the surrogate particle approach has the most uncertainties for an approximated infection risk. Leaving the aerosol generator the trajectories of the particles in the room are changing into omnidirectional motions, which might cause statistically based agglomeration. Depending on the concentration and the particle sizes, the number of particles agglomerating is increasing due to its probability of a collision. The intensity of this effect should be discussed in the extended results of figure 4, which lead to figure 11.

**Fig. 11.**
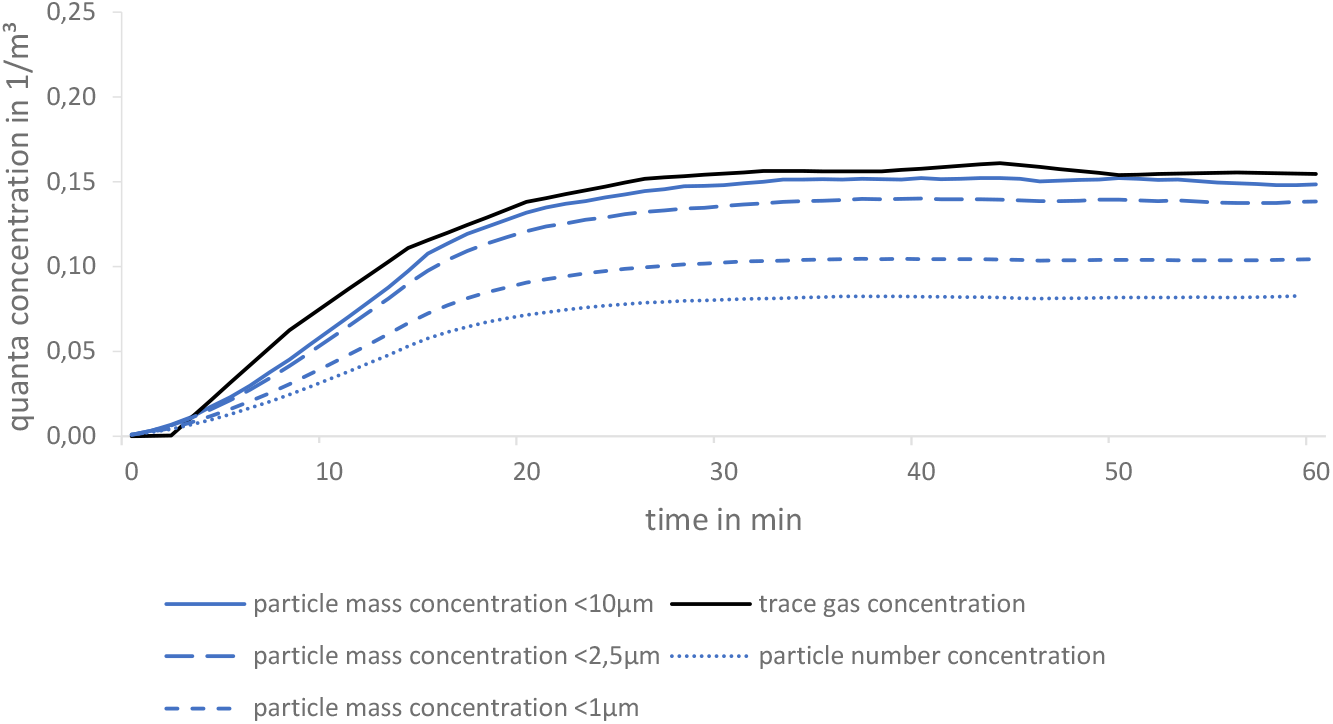
comparison of quanta concentrations based on particle mass, particle number and trace gas concentrations in a conference room

The assessment how well the quanta curves determined by surrogate particle method fit relatively the one by trace gas method is calculated as follows:

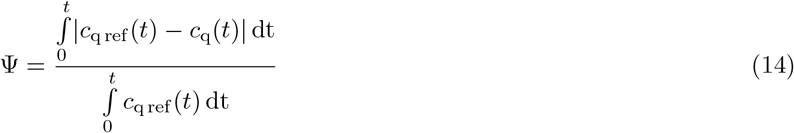

with Ψ, *c*_q ref_ as curve fit assessment and reference quanta concentration (trace gas method) respectively.

**Table 5.** curve fit assessment of different approaches

| approach | curve fit assessment $\Psi$ |
| --- | --- |
| trace gas (reference) | 0 % |
| particle number concentration | 48.7 % |
| particle mass concentration < 1 $\mu\text{m}$ | 35.0 % |
| particle mass concentration < 2.5 $\mu\text{m}$ | 13.3 % |
| particle mass concentration < 10 $\mu\text{m}$ | 5.6 % |

Agglomeration effects can not be considered using the particle number concentration, which results in Ψ = 48.7 %. For mass concentrations below 1 µm agglomeration might cause particles leaving the upper limit of the OPC’s detection range, which results in lower quanta concentrations compared to trace gas. For 2.5 µm this effect becomes a little smaller because of a lower amount of large particles. It is even negligible for 10 µm and its curve matches almost the trace gas course (Ψ = 5.6 %). To avoid underestimated infection risks it is highly recommended to apply mass concentrations < 10 µm.

Besides particle agglomeration further impacts on both methods might be caused by wall effects. Whereas trace gas should be reflected, liquid particles are assumed to be trapped at walls. This might be one possible explanation for a lower curve of particle mass compared to trace gas concentration (fig. 11).

In scenarios with both principles (filtering device and ventilation with outdoor air exchange), like an air purifier combined with a window or mechanical 24 ventilation, a new challenge arises. Outdoor air contains many particles of the same sizes as released by an aerosol generator, which means an OPC cannot separate between surrogate and outdoor particles. One pre-study in the project described in subsection 5.2 shows that a simultaneous outdoor SMPS measurement of particle size distribution provides a solution.

There are usually overlaps of the size distributions of outdoor particles and released ones. By taking into account only released particles outside this overlap, only these are considered by measuring instruments. It is worth mentioning that with this approach agglomeration effects can not be detected appropriately and therefore results in inaccuracies.

## 7 Conclusion

In order to estimate the infection risk of airborne indoor virus-transmissions either calculation models or measurements can be carried out. The implemented simplifications of these calculation models (e.g. ideal mixed ventilation) might deliver fast but inaccurate results for certain scenarios. Previous experimental studies examine the effects of ventilation measures in more detail. However, instead of transient and absolute considerations of viral loads they often regard relative statements of the infection risk.

Therefore, this study presents two experimental methods that are capable of determining a temporally and spatially resolved infection risk for different ventilation measures. Since particles entering from outside would falsify the particle measurements, only the trace gas method is suitable for ventilation systems with air exchange and for natural ventilation. However, an accurate assessment of air purifiers based on filtration is only applicable by the surrogate particle method, because trace gas is not filtered and therefore no device effect can be measured. For this reason, two different methods are essential.

Both methods are based on the theory that particles of relevant scales for infection procedures are completely airborne. The release of a controlled rate of either trace gas with a mass flow controller or particles with an aerosol generator allows a simulation of an infectious person releasing virus material. The measurement equipment includes a infrared spectrometer (trace gas method) or optical particle counters (surrogate particle method). For both approaches, the mathematical transfer of measured concentrations into infection risks is presented. In order to prove that the two methods are concordant, a comparison is essential. In an air visualisation laboratory with filtered outside air exchange both methods are executed simultaneously. In fact, they provide similar results. This allows a first-time reliable experimental comparison of ventilation systems, natural ventilation and air purifiers.

Besides the detailed explanations and the comparison of both methods, several aspects, which might influence the accuracy are, discussed. Two exemplary scenarios show how practical both methods are and how scalable this principle is. Even if the ventilation concept deviates significantly from mixed ventilation, infection risks can be determined. Besides a two person office, results of measurements performed in several classrooms are presented. Both scenarios highlight the value of experimental investigations with temporal and spatial resolutions for determining infection risks.

## Data Availability

All data produced in the present study are available upon reasonable request to the authors

## Acknowledgments

We would like to thank Michel Bauer for his scientific editing services.

## Declarations

- Data availability statement: Datasets generated/analysed during the current study are available from the corresponding author on reasonable request.
- Funding statement: The authors received no specific funding for this work.
- Conflict of interest disclosure: The authors declare no competing interests.
- Ethics approval statement: Not applicable.
- Patient consent statement: Not applicable.
- Permission to reproduce material from other sources: Not applicable.

## Notes

### Competing Interest Statement

The authors have declared no competing interest.

### Funding Statement

This study did not receive any funding

## References

[1] Lelieveld, J., Helleis, F., Borrmann, S., Cheng, Y., Drewnick, F., Haug, G., Klimach, T., Sciare, J., Su, H., Pöschl, U.: Model calculations of aerosol transmission and infection risk of covid-19 in indoor environments. International Journal of Environmental Research and Public Health 17(21) (2020). https://doi.org/10.3390/ijerph17218114

[2] Hussein, T., Löndahl, J., Thuresson, S., Alsved, M., Al-Hunaiti, A., Saksela, K., Aqel, H., Junninen, H., Mahura, A., Kulmala, M.: Indoor model simulation for covid-19 transport and exposure. International Journal of Environmental Research and Public Health 18(6) (2021). https://doi.org/10.3390/ijerph18062927

[3] Xu, C., Liu, W., Luo, X., Huang, X., Nielsen, P.V.: Prediction and control of aerosol transmission of sars-cov-2 in ventilated context: from source to receptor. Sustainable cities and society 76, 103416 (2022). https://doi.org/10.1016/j.scs.2021.103416

[4] Kennedy, M., Lee, S.J., Epstein, M.: Modeling aerosol transmission of sars-cov-2 in multi-room facility. Journal of loss prevention in the process industries 69, 104336 (2021). https://doi.org/10.1016/j.jlp.2020.104336

[5] Moreno, T., Pintó, R.M., Bosch, A., Moreno, N., Alastuey, A., Minguillón, M.C., Anfruns-Estrada, E., Guix, S., Fuentes, C., Buonanno, G., Stabile, L., Morawska, L., Querol, X.: Tracing surface and airborne sars-cov-2 rna inside public buses and subway trains. Environment international 147, 106326 (2021). https://doi.org/10.1016/j.envint.2020.106326

[6] Redder, C., Fieberg, C.: Sensitivity analysis of sars-cov-2 aerosol exposure. GMS hygiene and infection control 16, 28 (2021). https://doi.org/10.3205/dgkh000399

[7] Dinoi, A., Feltracco, M., Chirizzi, D., Trabucco, S., Conte, M., Gregoris, E., Barbaro, E., La Bella, G., Ciccarese, G., Belosi, F., La Salandra, G., Gambaro, A., Contini, D.: A review on measurements of sars-cov-2 genetic material in air in outdoor and indoor environments: Implication for airborne transmission. The Science of the total environment, 151137 (2021). https://doi.org/10.1016/j.scitotenv.2021.151137

[8] Li, Y., Qian, H., Hang, J., Chen, X., Cheng, P., Ling, H., Wang, S., Liang, P., Li, J., Xiao, S., Wei, J., Liu, L., Cowling, B.J., Kang, M.: Probable airborne transmission of sars-cov-2 in a poorly ventilated restaurant. Building and environment 196, 107788 (2021). https://doi.org/10.1016/j.buildenv.2021.107788

[9] Curtius, J., Granzin, M., Schrod, J.: Testing mobile air purifiers in a school classroom: Reducing the airborne transmission risk for sars-cov-2 (2020). https://doi.org/10.1101/2020.10.02.20205633

[10] Keene, C.H.: Airborne contagion and air hygiene. william firth wells. Journal of School Health 25(9), 249 (1955). https://doi.org/10.1111/j.1746-1561.1955.tb08015.x

[11] Riley, E.C., Murphy, G., Riley, R.L.: Airborne spread of measles in a suburban elementary school. American journal of epidemiology 107(5), 421–432 (1978). https://doi.org/10.1093/oxfordjournals.aje.a112560

[12] Buonanno, G., Morawska, L., Stabile, L.: Quantitative assessment of the risk of airborne transmission of sars-cov-2 infection: Prospective and retrospective applications. Environment international 145, 106112 (2020). https://doi.org/10.1016/j.envint.2020.106112

[13] van Doremalen, N., Bushmaker, T., Morris, D.H., Holbrook, M.G., Gamble, A., Williamson, B.N., Tamin, A., Harcourt, J.L., Thornburg, N.J., Gerber, S.I., Lloyd-Smith, J.O., de Wit, E., Munster, V.J.: Aerosol and surface stability of sars-cov-2 as compared with sars-cov-1. The New England journal of medicine 382(16), 1564–1567 (2020). https://doi.org/10.1056/NEJMc2004973

[14] Sze To, G.N., Chao, C.Y.H.: Review and comparison between the wells-riley and dose-response approaches to risk assessment of infectious respiratory diseases. Indoor air 20(1), 2–16 (2010). https://doi.org/10.1111/j.1600-0668.2009.00621.x

[15] Dabisch, P., Schuit, M., Herzog, A., Beck, K., Wood, S., Krause, M., Miller, D., Weaver, W., Freeburger, D., Hooper, I., Green, B., Williams, G., Holland, B., Bohannon, J., Wahl, V., Yolitz, J., Hevey, M., Ratnesar-Shumate, S.: The influence of temperature, humidity, and simulated sunlight on the infectivity of sars-cov-2 in aerosols. Aerosol Science and Technology 55(2), 142–153 (2021). https://doi.org/10.1080/02786826.2020.1829536

[16] Fisk, W.J., Seppanen, O., Faulkner, D., Huang, J.: Economic benefits of an economizer system: Energy savings and reduced sick leave (2004)

[17] Sun, Z., Ge, S.S.: Switched Linear Systems: Control and Design. Communications and Control Engineering. Springer, London (2005). http://swbplus.bsz-bw.de/bsz117170089cov.htm

[18] Maas, A.: Experimentelle Quantifizierung des Luftwechsels bei Fensterlüftung. Dissertation, University of Kassel, Kassel (1995-10-30)

[19] Morawska, L., Johnson, G.R., Ristovski, Z.D., Hargreaves, M., Mengersen, K., Corbett, S., Chao, C.Y.H., Li, Y., Katoshevski, D.: Size distribution and sites of origin of droplets expelled from the human respiratory tract during expiratory activities. Journal of aerosol science 40(3), 256–269 (2009). https://doi.org/10.1016/j.jaerosci.2008.11.002

[20] Hartmann, A., Lange, J., Rotheudt, H., Kriegel, M.: Emissionsrate und Partikelgröße von Bioaerosolen beim Atmen, Sprechen und Husten. https://doi.org/10.14279/DEPOSITONCE-10332

[21] Michaelides, E.E., Crowe, C.T., Schwarzkopf, J.D. (eds.): Multiphase Flow Handbook, Second edition edn. Mechanical and aerospace engineering. CRC Press Taylor & Francis Group, Boca Raton and London and New York (2017)

[22] Adili, M.R., Schmidt, M.: Ventilation effectiveness of residential ventilation systems and its energy-saving potential. In: Sayigh, A. (ed.) Sustainable Building for a Cleaner Environment: Selected Papers from the World Renewable Energy Network’s Med Green Forum 2017, pp. 451–462. Springer International Publishing, Cham (2019). https://doi.org/10.1007/978-3-319-94595-838

[23] Ai, Z., Mak, C.M., Gao, N., Niu, J.: Tracer gas is a suitable surrogate of exhaled droplet nuclei for studying airborne transmission in the built environment. Building simulation 13, 489–496 (2020). https://doi.org/10

[24] Asadi, S., Cappa, C.D., Barreda, S., Wexler, A.S., Bouvier, N.M., Ristenpart, W.D.: Efficacy of masks and face coverings in controlling outward aerosol particle emission from expiratory activities. Scientific reports 10(1) (2020). https://doi.org/10.1038/s41598-020-72798-7

[25] Hill, W.C., Hull, M.S., MacCuspie, R.I.: Testing of commercial masks and respirators and cotton mask insert materials using sars-cov-2 virion-sized particulates: Comparison of ideal aerosol filtration efficiency versus fitted filtration efficiency. Nano letters 20(10), 7642–7647 (2020). https://doi.org/10.1021/acs.nanolett.0c03182

[26] Palas GmbH: Datasheet DEHS, Karlsruhe (2021). https://palas.de/en/ product/download/dehs/datasheet/pdf Accessed 01.12.2021

[27] Bivolarova, M., Ondráček, J., Melikov, A., Ždímal, V.: A comparison between tracer gas and aerosol particles distribution indoors: The impact of ventilation rate, interaction of airflows, and presence of objects. Indoor air 27(6), 1201–1212 (2017). https://doi.org/10.1111/ina.12388

[28] DIN Deutsches Institut für Normung e. V.: DIN EN 16798-1 Energy performance of buildings – Ventilation for buildings –: Part 1: Indoor environmental input parameters for design and assessment of energy. Beuth Verlag GmbH (2021)

[29] Haycock, G.B., Schwartz, G.J., Wisotsky, D.H.: Geometric method for measuring body surface area: A height-weight formula validated in infants, children, and adults. The Journal of Pediatrics 93(1), 62–66 (1978). https://doi.org/10.1016/s0022-3476(78)80601-5

[30] Peng, Z., Pineda Rojas, A.L., Kropff, E., Bahnfleth, W., Buonanno, G., Dancer, S.J., Kurnitski, J., Li, Y., Loomans, M.G.L.C., Marr, L.C., Morawska, L., Nazaroff, W., Noakes, C., Querol, X., Sekhar, C., Tellier, R., Greenhalgh, T., Bourouiba, L., Boerstra, A., Tang, J.W., Miller, S.L., Jimenez, J.L.: Practical Indicators for Risk of Airborne Transmission in Shared Indoor Environments and Their Application to COVID-19 Outbreaks, (2021). https://doi.org/10.1101/2021.04.21.21255898

